# Influence of Information Access on Organ Donation: A Questionnaire-Based Cross-Sectional Study

**DOI:** 10.1101/2024.07.08.24310086

**Authors:** Guillermo A. Costaguta, Andrea Romero, Alejandro C. Costaguta

**Affiliations:** Servicio de Gastroenterología, Hepatología y Nutrición Pediátricas, CHU Sainte-Justine. 3175 Ch. de la Côte-Sainte-Catherine (H3T 1C5). Montréal, Canada; Gastroenterology Department of Sanatorio de Niños de Rosario, Alvear 863, CP 2000. Rosario, Argentina; Liver Transplant Unit of the Sanatorio de Niños de Rosario, Alvear 863, CP 2000. Rosario, Argentina

**Author notes:** **Correspondign author:** Guillermo Costaguta, MD, Division of gastroenterology, hepatology and nutrition CHU Sainte-Justine, 3175 Ch. de la Côte-Sainte-Catherine (H3T 1C5), Montréal, Quebec, **Working **(int 5295), **Personal **.

**Keywords:** organ donation – transplant – information – education level

## Abstract

**Introduction:** Organ transplantation is the sole effective treatment for end-stage organ diseases. However, the availability of donor organs remains insufficient. This shortage is driven by several factors, with access to accurate information being the key determinant of an individual’s willingness to donate organs.

**Methods:** A cross-sectional study based on anonymous surveys conducted from January to December 2019, categorizing participants into healthcare professionals and non-healthcare individuals. Data included willingness to donate organs, reasons for refusal, age, education level, and understanding of brain death. Statistical significance was set at p<0.05.

**Results:** A total of 408 participants were included: 203 in the healthcare sector and 205 in the non-healthcare sector. Among healthcare professionals, 90% were willing to donate organs compared to 43% in the non-healthcare group (p<0.001). Non-healthcare respondents refused due to the fear of being alive during organ removal (74%), concerns about reduced emergency care (21%), and religious beliefs (5%). Despite these concerns, 88% acknowledged that organ donation saves lives and 95% recognized the gap between organ supply and demand. No significant differences in education levels were found between donors and non-donors, but healthcare professionals had a significantly better understanding of brain death (p<0.001). All respondents indicated that they would accept a donated organ, if needed.

**Conclusion:** Healthcare professionals are more inclined to be organ donors than are those outside the field. Misunderstandings among non-healthcare individuals contributed to higher refusal rates. Tailored awareness campaigns and educational programs could rectify these misconceptions, potentially improving donation rates and mitigating organ shortage crises.

## INTRODUCTION

The concept of transplanting organs or tissues from a deceased individual to improve another’s health has fascinated physicians for centuries. One of the earliest recorded instances is The Miracle of Saints Cosmas and Damian in 348 AD. [1] However, it wasn’t until 1901 that Alexis Carrel described the first vascular suture techniques, marking a significant advancement in making transplantation a viable therapeutic option. [2] Over the following years, the discovery and investigation of immunological rejection emerged as the main obstacle to successful organ transplantation. A deeper understanding of the major histocompatibility complex (MHC) and human leukocyte antigen (HLA), primarily due to the contributions of Terasaki, Calne, and Starzl, provided crucial insights into overcoming this challenge. [3, 4] The development of cyclosporin as an immunosuppressant in the late 1970s finally established organ transplantation as an effective treatment. [4] The exponential increase in organ demand led to the foundation of the Centro Único Coordinador de Ablación e Implantes (CUCAI) in Argentina in 1977, a national institution dedicated to transplant coordination. In 1990, CUCAI evolved into the Instituto Nacional Central Único Coordinador de Ablación e Implantes (INCUCAI), a self-sufficient institution under the National Health Ministry.

Organ transplantation can involve two types of donors: living-related donors, suitable for organs such as kidneys, liver, or intestines, and deceased donors, from whom most transplanted organs are sourced. For deceased donation to be possible, the individual must be in a state called brain death. [5, 6] In recent years, “donation after cardiac death” (DCD) [7, 8] has gained popularity, though it remains infrequent in Argentina and is primarily used for tissue donation. [9, 10] However, the concept of brain death remains misunderstood by the general population, often confused with coma or a vegetative state. [11, 12]

Organ donation is an altruistic and voluntary process, where an individual expresses their wish to be a donor before their death, or their close relatives make the decision afterward. Although many laws in Argentina aim to increase the number of donors, the family’s final decision is always respected. Therefore, it is crucial for the population to understand the meaning of brain death, the process of organ donation, and the laws regulating it to ensure trust in the healthcare system and increase donor supply. [13, 14] (Figure 1) Various factors influence a person’s willingness to be a donor. However, access to adequate and relevant information is believed to be of paramount importance. This study aims to investigate whether being part of the healthcare system and having access to updated information affects the willingness to be an organ donor.

**FIGURE 1:**
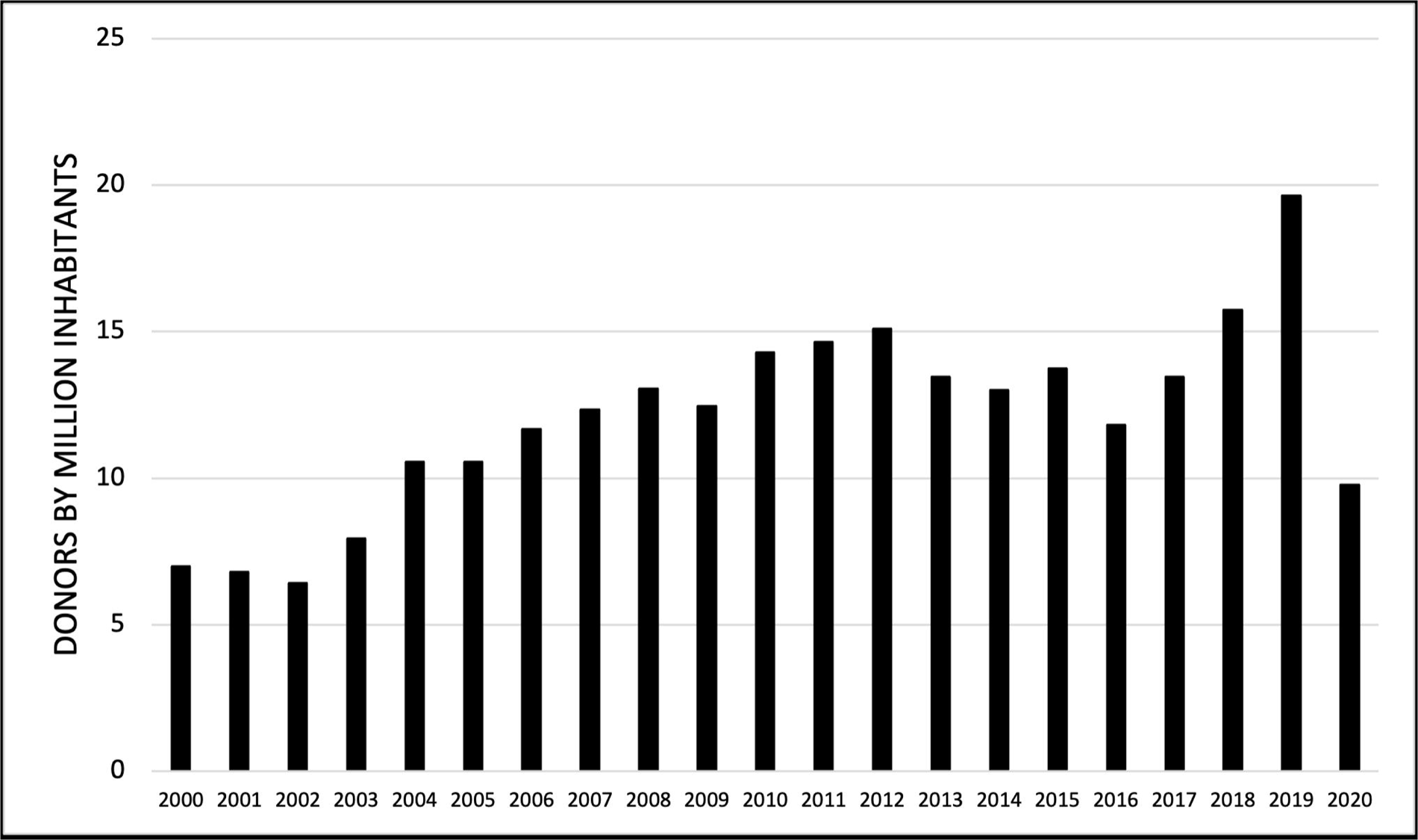
Donation rate by million inhabitants. Adapted from: SINTRA / CRESI. Memoria 2020: Procuración y Trasplante de Órganos, Tejidos y Células en Argentina. Ciudad Autónoma de Buenos Aires, 2021. [Consulted: November the 20th 2022]. Available in: https://bancos.salud.gob.ar/sites/default/files/2021-12/memoria-incucai-2020.pdf

**TABLE 1:** Population characteristics.

|  | **N-HS (n 205)** | **HS (n 203)** | **p** |
| --- | --- | --- | --- |
| **Donors** | 88 (43 %) | 182 (90 %) | **<0.001** |
| Sex M/F | 104/101 | 101/102 | 1 |
| Education |  |  |  |
| Primary/Secondary | 128 | 98 |  |
| **Tertiary/University** | 77 | 105 | **0.005** |
| Age |  |  |  |
| 18 to 29 years old | 82 | 73 | 0.42 |
| 30 to 49 years old | 70 | 70 | 1 |
| 50 to 65 years old | 53 | 60 | 0.5 |
| **Brain death** | 67 | 197 | **<0.001** |
| **Law of Presumed Donor** | 8 | 131 | **<0.001** |
| ¿**Would you accept an organ?** | 205 | 203 | **<0.001** |

*Statistically significant different variables are highlighted in bold*

## METHODS

A transversal descriptive study was conducted using anonymous surveys with multiple-choice questions from January to December 2019. Two populations were defined for the study: the healthcare system (HS) group, which included students from the Faculty of Medical Sciences at the Universidad Nacional de Rosario and staff from the Sanatorio de Niños in Rosario, and the non-healthcare (N-HS) group, which consisted of randomly selected individuals from Rosario city, ensuring the exclusion of anyone working in the healthcare system. This division was intended to represent the varying levels of access to quality information.

Participants included individuals older than 18 years who voluntarily agreed to participate and were mentally capable of doing so. Individuals older than 65 years were excluded, as they would not be considered potential donors.

The variables studied were age, sex, highest educational level attained, willingness to be an organ donor, reasons for refusal, ability to define brain death, and knowledge of the concept of “presumed donor.” The latter refers to the Argentinian law that considers every citizen older than 18 years a donor unless they explicitly opt out. Age was categorized into three ranges: 18–29, 30–49, and 50–65 years. Educational level was divided into primary/secondary and tertiary/university levels.

Data were analyzed using GraphPad Prism 9. Independent discrete variables were compared using the χ² test, Odds Ratio (OR), and 95% Confidence Interval (CI). Statistical significance was set at p < 0.05.

Ethics committee approval was obtained from the Sanatorio de Niños de Rosario, provided that informed consent was obtained from all participants.

## RESULTS

A total of 408 individuals from the city of Rosario, Santa Fe were surveyed, with 203 participants in the HS group and 205 in the N-HS group. The populations were similar except for the education level, which was higher in the HS group (p = 0.005).

Overall, 66% (n=270) of the entire population expressed willingness to be organ donors. However, significant differences were observed between the groups: 90% of the HS group (n=182) were willing to be donors, compared to only 43% of the N-HS group (n=88) (OR 11.52, CI 6.8–19.6, p<0.001). In the N-HS group, the primary reasons for refusal were concerns about being alive at the time of ablation (74%), doubts about treatment limitations in emergencies (21%), and religious beliefs (5%). Despite this, 88% (n=180) of the N-HS group believed that organ donation saves lives, and 95% (n=195) believed that organ supply is smaller than demand.

While differences in formal education levels were noted between the two groups, these differences were not significant when analyzed independently between donors and non-donors (OR 0.71, CI 0.47–1.8, p = 0.11). Notably, 97% of individuals in the HS group could define brain death (mainly regarding its irreversibility) and differentiate it from coma, whereas only 33% of the N-HS group could do so (OR 5.3, CI 3.5–8.1, p<0.0001). Within the HS group, a higher education level was directly correlated with the likelihood of being a donor (OR 0.34, CI 0.12–0.9, p = 0.03), a trend not observed in the N-HS group (OR 1.4, CI 0.79–2.5, p = 0.25). Regarding the knowledge of the “presumed donor” concept, 65% of the HS group were aware of the definition, compared to only 4% of the N-HS group (OR 44.8, CI 20.9–96, p<0.001).

Age also influenced the willingness to be an organ donor. Those aged 18-29 years were more likely to express willingness to donate their organs than those aged 30-49 years (OR 4.77) and those aged 50-65 years (OR 2.04, p<0.001). Additionally, individuals aged 30-49 years were more willing to donate compared to those aged 50-65 years (OR 1.89, p<0.001). There were no significant differences between the sexes of donors and non-donors (OR 0.68, CI 0.45–1, p = 0.07).

Of those surveyed, 100% indicated they would accept a donated organ, regardless of whether they were organ donors themselves (p<0.001).

## DISCUSION

In this study we aimed to assess differences in organ donor likelihood between the HS and N-HS groups and explore reasons for refusal. Our primary objective was to evaluate the impact of information access, using this division as a surrogate. This interest arose from the necessity to identify variables influencing willingness to donate organs, with the goal of informing strategies to increase donor numbers.

In our surveyed population, 66% expressed willingness to be organ donors, a figure notably lower than the 88% reported by Coelho et al. [15], but more aligned with rates around 75% found by Shahbazian et al. [16] and Truog et al. [17] Within the HS group, acceptance was higher at around 90%, consistent with findings among physicians and medical students. [18–22] Interestingly, Terada et al. [23] found less support for organ donation after brain death among nurses, a sentiment also observed among nursing students in a study by a Chinese team. [24] This suggests that while education and information access are important, additional factors influence attitudes towards organ donation. [25] In contrast, acceptance rates in the N-HS group in our study were similar to those reported by other studies. [18, 26–28]

While we did not observe a linear correlation between educational level and the likelihood of being an organ donor in our population, some studies have reported such associations. [15, 16] Conversely, Salmani et al. [19] found no significant differences based on education. However, when we analyzed education level within each group independently, we found a direct correlation only in the HS group. This finding, though not extensively studied, suggests that targeted education on organ donation increases acceptance rates [22, 23, 30–34]. For instance, McGlade et al. [25] reported an increase in willingness to donate from 35% to 45% following transplantation education, with 53% initiating family discussions on the topic. Similarly, Park et al. [35] noted increased individual willingness to donate organs but found no change in attitudes towards family member donations. These findings imply that higher education and better access to information facilitate understanding and acceptance of new concepts, contingent on individual interest. These insights are underscored by reasons for refusal among non-donors in our study. A significant proportion (75%) feared being alive during organ removal, while 21% worried about potential treatment limitations in emergencies due to organ donor status. These concerns often stem from misinformation or distrust in healthcare systems. [15, 27] When questioned about their views on organ donation, 88% of non-donors expressed belief that organ donation saves lives or improves quality of life, while 95% felt the current number of donors falls short of needs. Similar sentiments have been reported by other researchers [15, 21], with 88% acknowledging that organ transplantation enhances patients’ lives and enables normalcy.

In addition to information access, our study revealed that only 52% of the general population understood the irreversibility of brain death. Comparatively, 97% of the HS group correctly defined brain death, whereas only 33% of the N-HS group did so. These findings echo similar studies [32, 36, 37], which have also noted challenges in understanding brain death even among medical students [38, 39, 40] and transplant team members. [38, 39]

Age emerged as a significant factor influencing donation rates, with individuals aged 18-29 more likely to express willingness to donate compared to those aged 50-65. This aligns with previous research findings. [15, 27] While some studies suggest religion may influence donation willingness, this aspect wasn’t explored in our study, except for one participant citing religious reasons against donation. [41]

This study, involving a substantial number of participants with limited comparable literature, offers conclusions that may broadly reflect societal trends. However, grouping by healthcare and non-healthcare sectors could oversimplify subpopulations, potentially obscuring nuanced intragroup differences.

## CONCLUSIONS

Organ donation is indeed a multifaceted issue with varying beliefs across different segments of society. The disparity between organ availability and demand remains stark, leading to tragic outcomes where many individuals perish while awaiting transplantation. Understanding the factors influencing individuals’ decisions to become organ donors is crucial as it enables targeted interventions on modifiable factors. Access to accurate information and comprehensive education on organ donation appear pivotal in shaping public attitudes. Despite numerous awareness campaigns over recent decades, which have shown some positive effects, organ donation rates continue to fall short of optimal levels.

## Data availability

Data will be made available when properly justified in the interest of research by sending an email to the corresponding author.

## Authors contributions

Guillermo Costaguta: Conception and design of the study, acquisition, analysis, and data interpretation. Drafting of the work and critical revisions. Final approval of the version to be published. Accountability for all aspects of work.

Romero Andrea: conception and design of the work, acquisition, analysis, and data interpretation. Drafting of the work and critical revisions. Final approval of the version to be published. Accountability for all aspects of work.

Costaguta Alejandro: conception and design of the work, acquisition, analysis, and data interpretation. Drafting of the work and critical revisions. Final approval of the version to be published. Accountability for all aspects of work.

